# Risk factors for suicide and repeat self-harm: a cohort study of adults with hospital-presenting self-harm

**DOI:** 10.64898/2026.06.15.26355458

**Authors:** Oskar Flygare, Johan Bjureberg, John Wallert, Sabrina Doering, Ellinor Salander Renberg, Margda Waern, Bo Runeson

**Author notes:** Corresponding author: Oskar Flygare,. Centre for Psychiatry Research, Norra Stationsgatan 69, 113 64 Stockholm, Sweden. E-mail addresses: Oskar Flygare, Johan Bjureberg, John Wallert, Sabrina Doering, Ellinor Salander Renberg, Margda Waern, Bo Runeson.

## Abstract

**Background:** Previous self-harm elevates the risk of repeat self-harm and suicide, but the prognostic value of events and clinician observations around the index event is unclear. We evaluated established and exploratory risk factors for suicide and repeat self-harm among patients presenting to emergency psychiatric units after a suicide attempt or nonsuicidal self-injury (NSSI).

**Methods:** Multicentre cohort study in Sweden (n = 804). Outcomes were suicide and repeat self-harm at 1-year and 5-year follow-up, ascertained through linked national registers. Established risk factors included psychiatric diagnoses, prior suicidal behaviour, and sociodemographic characteristics; exploratory factors comprised past-week self-reported symptom changes and clinician observations. LASSO-regularised Cox regression models were fitted for established (n=21) and exploratory (n=11) risk factors.

**Results:** During five-year follow-up, 285 (35%) individuals had a new episode of self-harm and 41 (5%) died by suicide. No risk factors reached statistical significance for suicide, although male sex was retained after regularisation (1-year hazard ratio [HR] = 3.57 [95% CI 0-8.33]; 5-year HR = 2.5 [0.03-4.55]). Three established risk factors were significantly associated with repeat self-harm: psychiatric inpatient care in the three months before the index event (1-year HR = 1.85 [1.3-2.6]; 5-year HR = 1.72 [1.23-2.65]), previous suicide attempt (1-year HR = 2.01 [0.79-2.4]; 5-year HR = 2.19 [1.27-2.6]), and borderline personality disorder (1-year HR = 1.82 [1.13-3]; 5-year HR = 1.67 [0.14-2.75]). Among exploratory risk factors, clinician-observed hopelessness (1-year HR = 1.72 [1.1-2.3]; 5-year HR = 1.51 [1.03-1.91]) and personality disorder features (1-year HR = 1.48 [0.96-2.05]; 5-year HR = 1.47 [1.04-1.95]) were associated with repeat self-harm.

**Conclusions:** Risk factor profiles for repeat self-harm were consistent at 1 and 5 years. Beyond established risk factors, clinician-observed hopelessness and personality disorder features emerged as markers of risk, suggesting that qualitative clinician assessments may yield prognostic information not available from medical records alone.

## 1. Introduction

Suicide is a global public health problem, with 750,000 deaths each year, accounting for 1.3% of all deaths worldwide (Davis Weaver et al., 2025; WHO, 2021). Previous self-harm and use of violent methods to attempt suicide are among the strongest risk factors for subsequent suicide among individuals who are seen in hospitals after a suicide attempt (Fedyszyn et al., 2016; Runeson et al., 2010). Five years after an index suicide attempt, 1 in 20 patients die by suicide (Demesmaeker et al., 2022). Understanding the contributing factors for this elevated risk is therefore a research priority.

Several risk factors have been consistently linked to increased risk of suicide or reattempt. These include mental disorders (e.g., borderline personality disorder, depression, bipolar disorder, autism spectrum disorder and substance use disorder) (Chesney et al., 2014; Fazel & Runeson, 2020; Hirvikoski et al., 2019; Pemau et al., 2024), previous suicidal behaviour (Ribeiro et al., 2016; Runeson et al., 2017), poor physical health (Favril et al., 2022), access to lethal means (Mann & Michel, 2016), and a family history of suicide or suicidal behaviour (Qin et al., 2002). The link between socioeconomic status and suicide is less clear. Although life expectancy increases with higher income (Kinge et al., 2019), mortality rates due to external causes (including suicide) are higher among individuals with high socioeconomic position after adjusting for mental disorders (Chen et al., 2024). Negative life events, for example spousal suicide (Erlangsen et al., 2017), receiving a cancer diagnosis (Fang et al., 2012), and relationship difficulties, have been studied for more than 50 years (Paykel et al., 1975), but the evidence is weaker compared to stable risk factors such as psychiatric history (Fazel & Runeson, 2020). Recently, there has been an increased interest in evaluating dynamic changes in negative emotional states such as hopelessness and loneliness (Kleiman et al., 2023), and the authors of a comprehensive meta-analysis evaluating 50 years of research on risk factors for suicidal thoughts and behaviours highlighted a need for studies that measure changes in these emotions alongside stable risk factors (Franklin et al., 2017).

The impact of specific risk factors may also vary over time. For example, one study of 954 patients with depression found that risk factors differed between individuals who died by suicide within one year after discharge from psychiatric care compared to 2-10 years after discharge (Fawcett et al., 1990). A recent population-based study identified variations in the relative risk of 11 risk factors up to two years after discharge (Aaltonen et al., 2024). In ideation-to-action theories of suicide, suicide risk fluctuates as a result of interactions between stable risk factors (listed above) and short-term dynamic risk factors such as recent life events and changes in mood (Bryan & Rudd, 2016; Klonsky et al., 2018). Few prospective cohort studies, however, have combined the assessment of both stable and dynamic risk factors, or have been limited to individuals with affective disorders, whereas the population attending psychiatric units after a suicide attempt is more diverse.

### 1.1 Objective

Building on previous work in the field, the aims of the present study were to: (1) evaluate the most important risk factors for self-harm repetition and suicide, including established risk factors as well as exploratory risk factors based on past-week changes and clinician observations in connection with the index event; (2) examine whether these risk factors were stable across 1- and 5-year follow-up; and (3) evaluate the risk factors among individuals with major depressive disorder.

## 2. Methods

### 2.1 Study design

Data for this study were collected as part of a multicentre cohort study at three Swedish psychiatric departments in Stockholm, Gothenburg and Umeå, from April 2012 through April 2016. Ethical approval was obtained from the Göteborg Regional Ethics Committee (589-10, T034-12). Follow-up was done through medical records review and linked national registers one and five years after inclusion.

### 2.2 Participants

Patients 18 years or older presenting to the psychiatric emergency departments within a week after a self-harm event (with or without suicidal intent) were invited to participate in the study. Patients also needed to have a Swedish personal identity number since it was required for medical record and register follow-up. Exclusion criteria were confusion, pronounced aggression, vivid psychotic symptoms or not speaking Swedish, as the project involved a comprehensive psychiatric interview. If these serious symptoms subsided within a few days, patients were eligible for inclusion. Out of 1134 eligible patients, 804 (71%) accepted to participate and were included. At baseline, 666 participants (83% of the total sample) presented with a suicide attempt, defined in accordance with the Columbia Suicide Severity Rating Scale (C-SSRS) as an act of physical self-injury with at least some degree of intent to die (Lindh et al., 2019). In the remaining 138 participants (17%), the baseline self-injury was without intent to die and consequently defined as non-suicidal self-injury (NSSI).

### 2.3 Measures

The established risk factors were obtained from a comprehensive clinical interview conducted at baseline by mental health staff (psychiatrist, psychologist or psychiatric nurse). These included background information on age, sex (male, female), highest level of completed education (none, primary school, secondary school, university), unemployment (yes/no), whether the patient was living alone, and if any close relatives had died by suicide (parents, grandparents, siblings, children or partner). The MINI psychiatric interview was administered to assess current psychiatric diagnoses of depression, alcohol use disorder, substance use disorder and post-traumatic stress disorder (Sheehan et al., 1998). As personality disorders, attention-deficit/hyperactivity disorder (ADHD), and autism spectrum disorders were not included in this version of the MINI, these diagnoses were retrieved from medical charts at the time of the index episode. Patients also reported whether they had problems with chronic pain (yes/no). The C-SSRS was used to assess lifetime suicide attempts and/or previous NSSI (yes/no, described above) (Posner et al., 2011). The Karolinska Interpersonal Violence Scale (KIVS) was administered to assess whether the patient had been violent as a child or adult (yes if violent as either child or adult, otherwise no) (Jokinen et al., 2010). The index self-harm events were classified as impulsive if the individual responded that they had made no active preparations and no premeditation (items 6 and 15 of Beck’s Suicidal Intent Scale rated 0) (Baca-Garcia et al., 2005; Lindh et al., 2019). Finally, clinicians also assessed the primary method of self-harm at the index event (poisoning or other method). The total number of these established risk factors was 21.

In addition to the established risk factors listed above, we also evaluated new exploratory risk factors of past-week changes in psychiatric symptoms and clinician observations during the baseline clinical interview. Patients indicated if they had experienced past-week changes in restlessness, anxiety, depressive symptoms, hopelessness, relationship problems, or alcohol intake (all reported as increased or unchanged). The clinician also made observations regarding patient behaviour during the interview: whether they appeared restless, anxious, expressed hopelessness, were sincere in their responses, or showed signs of a personality disorder (e.g., showed impulsivity, aggression, narcissism or antisocial tendencies), all coded as present or not present. There were 11 of these exploratory risk factors in total.

### 2.4 Outcomes

Suicide and repeat self-harm were evaluated at 1 year and 5 years after the index event using data from linked national registers (Ludvigsson et al., 2016). Death by suicide was defined as death from intentional self-harm (ICD-10 codes X60-X84) or events of undetermined intent (Y10-Y34) in the *National Cause of Death Register*; self-inflicted deaths of undetermined intent were included to avoid underestimation of suicides (Neeleman & Wessely, 1997; Runeson et al., 2010). Repeat self-harm was defined as either death by suicide or a visit to outpatient or inpatient health care due to deliberate self-harm or events of undetermined intent in the *National Patient Register*, using the same ICD-10 codes.

### 2.5 Statistical analyses

In each set of established and exploratory risk factors, the selected risk factors were pre-processed by only keeping those with sufficient variance (i.e., at least 5% of participants indicating a certain response), independence from other variables (i.e., correlation ≤0.8), and less than 30% missing observations. Missing data in the risk factor variables were imputed using bagged trees (Kuhn & Johnson, 2013) and all continuous variables were centred in preparation for statistical modelling.

We fitted eight penalised Cox regression models for each combination of outcomes (suicide, repeat self-harm), follow-up points (1 year, 5 years) and risk factor sets (established, exploratory). Regularisation was performed using the least absolute shrinkage and selection operator (LASSO) with 5-fold cross-validation to select the optimal *»* value, evaluating model fit using the Harrell C-index (Harrell et al., 1982; Simon et al., 2011). Coefficients, *p*-values and confidence intervals from the predictors remaining after regularisation were obtained using methods developed for selective inference, which can be used to provide accurate estimates after LASSO regularisation (Taylor & Tibshirani, 2018; Taylor & Tibshirani, 2015). Predictors with a *p*-value lower than 0.05 were considered statistically significant and were visualised using adjusted survival curves, showing the probability of being event-free in the presence or absence of each risk factor (fitted separately) over the 5-year follow-up period (Denz et al., 2023; Hernán, 2010). Crude hazard ratios obtained from univariate Cox models were also reported for comparison. In suicide models, follow-up started at the index event and participants were right-censored at non-suicide death or administrative end of follow-up. In the repeat self-harm models, the outcome also included non-fatal self-harm events in addition to death by suicide, and only the first self-harm event was analysed. All statistical analyses were conducted using R (version 4.6.0) (R Core Team, 2026).

## 3. Results

Baseline characteristics of the sample are shown in Table 1. A total of 41 (5.1%) participants died by suicide, 19 within the first year (incidence rate per 100 000 person-years [IR] = 2416 [95% CI 1541, 3787]) and 22 in subsequent years (IR = 742 [489, 1127]). There were 285 (35.4%) individuals with repeat self-harm, of which 178 (IR = 26 447 [22 833, 30 632]) had a first repetition within the first year and 107 (IR = 4695 [3884, 5674]) in subsequent years.

**Table 1:** Baseline characteristics of the study sample (N = 804)

|  | No. (%) | Missing data, n (%) |
| --- | --- | --- |
| **Established risk factors** |  |  |
| Sociodemographic characteristics |  |  |
| Sex |  |  |
| Female | 541 (67.3) |  |
| Male | 263 (32.7) |  |
| Age-group |  |  |
| 18-29 | 356 (44.3) |  |
| 30-44 | 186 (23.1) |  |
| 45-64 | 180 (22.4) |  |
| 65+ | 82 (10.2) |  |
| Highest attained education |  | 1 (0.1) |
| None | 16 (2.0) |  |
| Primary | 211 (26.2) |  |
| Secondary | 402 (50) |  |
| University | 174 (21.6) |  |
| Currently unemployed | 134 (16.7) |  |
| Living alone | 424 (52.7) |  |
| Suicide of family member | 89 (11.1) |  |
| *Characteristics of the index event* |  |  |
| Suicide attempt at index | 666 (82.8) |  |
| Impulsive suicide attempt at index | 309 (38.4) | 18 (2.2) |
| NSSI at index | 138 (17.2) |  |
| Violent method used at index | 196 (24.4) |  |
| *History of suicidal behavior* |  |  |
| Previous suicide attempt | 552 (68.7) |  |
| Previous NSSI | 464 (57.5) | 13 (1.6) |
| Ever violent | 412 (51.2) | 26 (3.2) |
| Psychiatric inpatient care in past 3 months | 231 (28.7) | 1 (0.1) |
| *Current diagnoses* |  |  |
| Major depressive disorder | 550 (68.4) | 40 (5.0) |
| Borderline personality disorder | 138 (17.2) | 8 (1.0) |
| Alcohol use disorder | 39 (4.9) | 54 (6.7) |
| Substance use disorder | 11 (1.4) | 48 (6.0) |
| PTSD | 153 (19.0) | 47 (5.8) |
| ADHD | 72 (9.0) | 8 (1.0) |
| ASD | 54 (6.7) | 8 (1.0) |
| Chronic pain | 189 (23.5) |  |
| **Exploratory risk factors** |  |  |
| *Past-week changes* |  |  |
| Restlessness | 578 (71.9) | 2 (0.2) |
| Anxiety | 674 (83.8) | 1 (0.1) |
| Hopelessness | 735 (91.4) | 1 (0.1) |
| Relationship problems | 526 (65.4) | 1 (0.1) |
| Depressive symptoms | 609 (75.7) | 45 (5.6) |
| Alcohol use | 149 (18.5) | 44 (5.5) |
| *Clinician observations during assessment* |  |  |
| Restless | 198 (24.6) | 25 (3.1) |
| Anxious | 237 (29.5) | 22 (2.7) |
| Hopelessness | 419 (52.1) | 22 (2.7) |
| Appears sincere | 732 (91.0) | 36 (4.5) |
| Personality disorder indicators* | 300 (37.3) | 77 (9.6) |
| Abbreviations: ADHD, Attention-deficit hyperactivity disorder; ASD, Autism-spectrum disorder; NSSI, non-suicidal self-injury; PTSD, Post-traumatic stress disorder. |  |  |
| * The clinician assessed whether the patient showed impulsivity, aggression, narcissistic or antisocial tendencies during the baseline clinical interview. |  |  |

### 3.1 Risk factors for suicide

There were no statistically significant risk factors of suicide at the 1-year follow-up or 5-year follow-up in the penalised models, although the models retained the coefficient for sex with an elevated risk for males (1-year HR = 3.57 [0 to 8.33], p=0.469; 5-year HR = 2.5 [0.03 to 4.55], p=0.469). Table 2 shows an overview of penalised and crude associations between statistically significant risk factors and the outcomes of interest. See supplementary material for a table including the results for all evaluated risk factors.

**Table 2.** Penalized and crude estimates of associations between risk factors and outcomes. HR (95% CI)

|  |  |  |  |  |
| --- | --- | --- | --- | --- |
| Risk factor | Penalized estimate* | Crude estimate† | Penalized estimate* | Crude estimate† |
|  | **Suicide <1y** |  | **Suicide 1-5y** |  |
| Male sex | 3.57 (0 to 8.33) | 3.57 (1.41 to 9.09) | 2.5 (0.03 to 4.55) | 2.5 (1.37 to 4.76) |
|  | **Repeat self-harm <1y** |  | **Repeat self-harm 1-5y** |  |
| Psychiatric inpatient care | 1.85 (1.3 to 2.6) | 2.61 (1.97 to 3.48) | 1.72 (1.23 to 2.65) | 2.4 (1.9 to 3.04) |
| Previous suicide attempt | 2.01 (0.79 to 2.4) | 2.93 (1.96 to 4.37) | 2.19 (1.27 to 2.6) | 2.98 (2.16 to 4.11) |
| Borderline personality disorder | 1.82 (1.13 to 3) | 2.97 (2.2 to 4.01) | 1.67 (0.14 to 2.75) | 2.69 (2.09 to 3.48) |
| Hopelessness§ | 1.72 (1.3 to 2.3) | 1.72 (1.28 to 2.32) | 1.51 (1.03 to 1.91) | 1.6 (1.26 to 2.03) |
| Personality disorder features§ | 1.48 (0.96 to 2.05) | 1.59 (1.2 to 2.11) | 1.47 (1.04 to 1.95) | 1.66 (1.32 to 2.1) |
| Abbreviations: CI, confidence interval; HR, hazard ratio. |  |  |  |  |
| * Coefficient estimate from LASSO-regularized model. The models included all risk factors in the sets of established and exploratory risk factors, respectively. |  |  |  |  |
| † Coefficient estimate from Cox proportional hazards model with each risk factor entered separately. |  |  |  |  |
| § Risk factor from the set of exploratory risk factors. |  |  |  |  |

### 3.2 Risk factors for repeat self-harm

Individuals with inpatient psychiatric care in the three months prior to the index event (1-year HR = 1.85 [1.3 to 2.6], p<0.01; 5-year HR = 1.72 [1.23 to 2.65], p<0.01), a previous suicide attempt (1-year HR = 2.01 [0.79 to 2.4], p=0.075; 5-year HR = 2.19 [1.27 to 2.6], p<0.01), and a diagnosis of borderline personality disorder (1-year HR = 1.82 [1.13 to 3], p=0.011; 5-year HR = 1.67 [0.14 to 2.75], p=0.358) had an elevated risk of repeat self-harm and statistically significant associations at one or both follow-up points. In addition to these three established risk factors, the models evaluating exploratory risk factors further identified hopelessness during the clinical interview (1-year HR = 1.72 [1.1 to 2.3], p=0.011; 5-year HR = 1.51 [1.03 to 1.91], p=0.018) and personality disorder features (1-year HR = 1.48 [0.96 to 2.05], p=0.034; 5-year HR = 1.47 [1.04 to 1.95], p=0.016) as statistically significant risk factors associated with increased risk. Adjusted survival curves for these risk factors are shown in Figure 1.

### 3.3 Risk factors among patients with major depressive disorder

The results were largely similar in subgroup analyses of risk factors among the 550 individuals with a current diagnosis of major depressive disorder. There were no statistically significant predictors of suicide, with the exception of male sex at 5-year follow-up, HR = 3.33 [0.83 to 7.14], p=0.04.

The risk factors with statistically significant associations with repeat self-harm at at least one follow-up point were once again recent inpatient psychiatric care (1-year HR = 1.62 [0.76 to 4.13], p=0.084; 5-year HR = 1.55 [0.96 to 2.19], p=0.034), a previous suicide attempt (1-year HR = 1.86 [0.9 to 3.84], p=0.043; 5-year HR = 2.03 [0.93 to 2.45], p=0.037), and a diagnosis of borderline personality disorder (1-year HR = 2.1 [1.42 to 18.63], p<0.01; 5-year HR = 1.55 [0.42 to 2.49], p=0.221). In addition, previous NSSI (1-year HR = 1.56 [0.99 to 34.01], p=0.026; 5-year HR = 1.68 [0.98 to 2.27], p=0.03) and a diagnosis of autism spectrum disorder (1-year HR = 3.33 [1.84 to 5.88], p<0.01) were associated with an elevated risk of repeat self-harm, although autism spectrum disorder was removed during LASSO-regularisation at the 5-year follow-up. Among the exploratory risk factors for repeat self-harm, hopelessness (1-year HR = 1.64 [0.72 to 2.25], p=0.107; 5-year HR = 1.48 [0.36 to 1.87], p=0.348) and personality disorder features (1-year HR = 1.44 [0.25 to 1.92], p=0.427; 5-year HR = 1.51 [0.74 to 1.97], p=0.115) were retained in the models after LASSO-regularisation with effects of similar magnitude to the full sample, but these associations did not reach statistical significance in the smaller subsample.

## 4. Discussion

This cohort study evaluated risk factors for suicide and self-harm repetition among 804 individuals seen at a psychiatric emergency unit after a suicide attempt or NSSI, and also compared whether risk factor profiles differed between 1-year and 5-year follow-up. Male sex was associated with a non-significant increase in suicide risk across both follow-up periods. For repeat self-harm, inpatient psychiatric care in the three months preceding the index event, previous suicide attempt, and borderline personality disorder emerged as important risk factors. In addition, we found that clinician observations of hopelessness and personality disorder features during the clinical interview were associated with elevated risk of repeat self-harm. These findings were consistent across both the 1-year and 5-year follow-up points and in a subsample of individuals with major depressive disorder.

The persistence of the identified risk factors for repeat self-harm at both follow-up points underscores that they reflect stable vulnerability rather than time-limited indicators of acute crisis. This consistency contrasts with some reports suggesting that the relative importance of risk factors may shift over time in clinical populations (Aaltonen et al., 2024; Fawcett et al., 1990), and may indicate that among individuals seen at emergency departments after self-harm, a core set of factors characterise those at highest long-term risk. The most important risk factors for repeat self-harm (prior psychiatric hospitalisation, history of previous suicide attempt, borderline personality disorder) are established risk factors in the field (Franklin et al., 2017), and are consistently linked to elevated risk in cohort studies of hospital-presenting self-harm (Cully et al., 2021; Cully et al., 2024) as well as register-based studies (Bøe et al., 2022).

In the day-to-day assessment of suicidal patients, clinicians are most often concerned about fluctuating risk factors such as ongoing crises, whereas research studies tend to evaluate static risk factors such as medical history since that information can easily be collected at scale (Nguyen et al., 2025). Our findings that clinician-observed hopelessness and personality disorder features are associated with increased risk of self-harm repetition highlight the value of a comprehensive psychiatric assessment of patients presenting with self-harm. These types of observations have the potential to improve the accuracy of risk prediction models, compared to models using electronic health records data alone (Nock et al., 2022). Suicide prevention guidelines also recommend a thorough assessment of the acute context that aids clinicians in formulating a shared understanding of the current episode with the patient, building a therapeutic alliance, and providing interventions such as safety planning (Franklin et al., 2017; NICE, 2022; Stanley et al., 2018). We also note that none of the events or symptom changes reported over the past week (including relationship problems, increased alcohol use, and worsening depressive symptoms) were statistically significant predictors of repeat self-harm in this sample. Future research should therefore aim to identify which dynamic risk factors meaningfully improve risk prediction and warrant assessment in studies using frequent data collection methods such as ecological momentary assessment (Kleiman et al., 2023; Wang et al., 2021).

Several limitations should be mentioned. The sample size did not allow for analysis of risk factors in sub-groups by age and sex. This should be taken into consideration when interpreting the results since the prevalence and importance of specific risk factors likely vary by age and sex (Johansson et al., 2025). Related to this, the study was not of sufficient size to detect statistically significant risk factors for suicide, although the non-significant finding that males were three times more likely than women to die by suicide after the index event was consistent across models and of the expected magnitude (Geulayov et al., 2019). The generalisability of our findings is also limited to emergency psychiatric settings, as risk factors in the general population or other clinical contexts may differ. The strengths of this study include the well-characterised clinical cohort which enabled a combined analysis of both well-established risk factors and clinical observations, which has often been lacking in previous studies. In addition, follow-up was done through linked national registers with minimal data loss.

### 4.1 Conclusions

In this study of 804 individuals seen at psychiatric emergency departments after an index self-harm event, recent inpatient psychiatric care, previous suicide attempt, and a diagnosis of borderline personality disorder were associated with repeat self-harm. In addition, clinician-made observations of hopelessness and personality disorder features were also associated with an elevated risk of repeat self-harm. The findings were consistent at 1-year and 5-year follow-ups, and in a subsample of patients with major depressive disorder. Our results underscore the value of a comprehensive psychiatric assessment for individuals presenting with self-harm, incorporating observations made by the clinician alongside well-known markers of risk.

## Supporting information

STROBE checklist

## Additional information

## Acknowledgements

The authors are grateful to the study participants and clinical staff who contributed to this research.

## CRediT authorship contribution statement

*Concept and design*: Flygare, Bjureberg, Wallert, Runeson. *Acquisition, analysis, or interpretation of data*: All authors. *Drafting of the manuscript*: Flygare, Runeson. *Critical revision of the manuscript for important intellectual content*: All authors. *Statistical analysis*: Flygare. *Obtained funding*: Waern, Salander Renberg, Runeson. *Administrative, technical, or material support*: Waern, Salander Renberg, Runeson. *Supervision*: Bjureberg, Runeson.

## Data availability statement

Data used for analyses contain sensitive personal identifying information and are not publicly available as data sharing was not part of the written informed consent. Data could be made available from the corresponding author upon reasonable request following approval from the Swedish Ethical Review Authority.

---

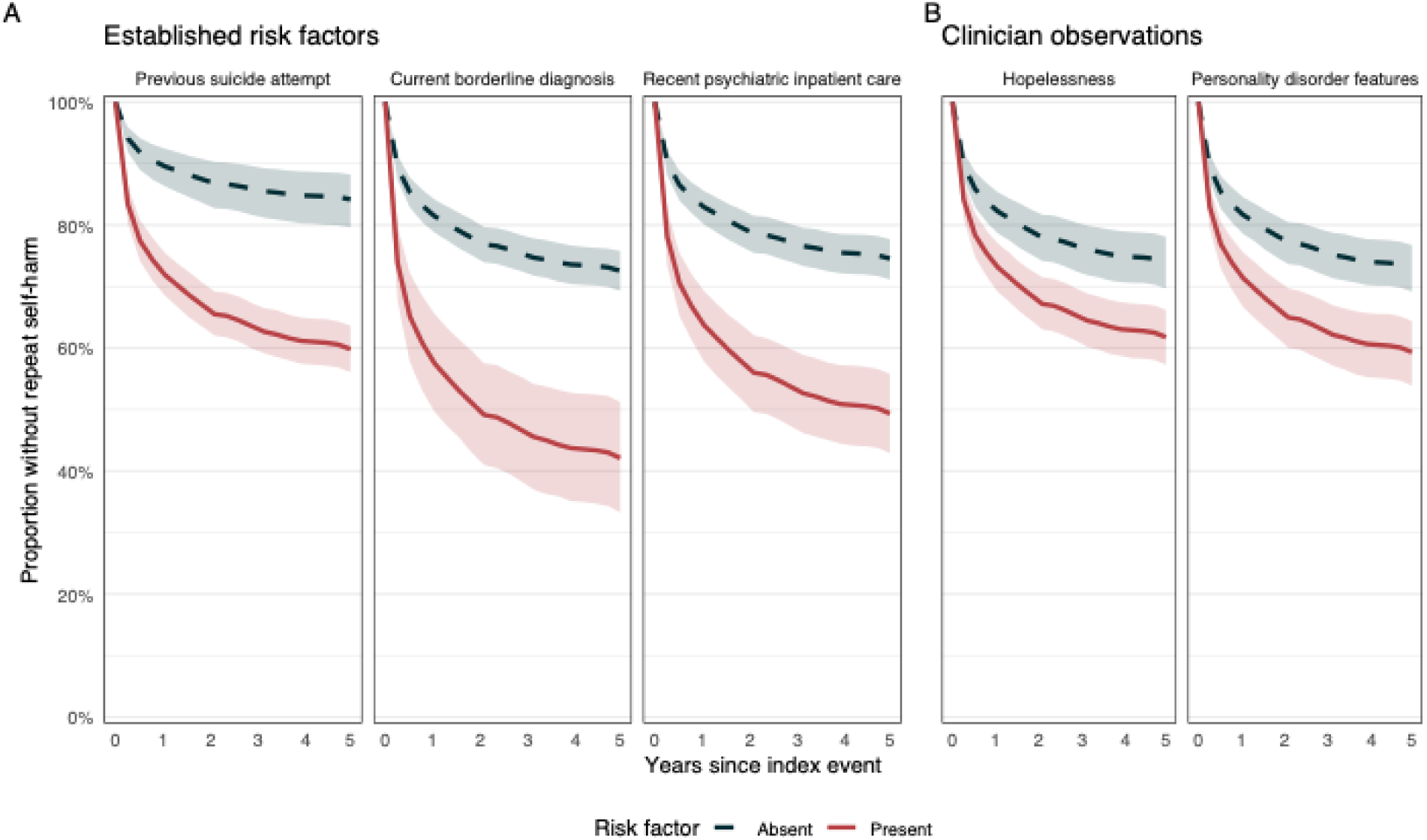

